# Longitudinal Dynamics and Pluripotentiality of Polysymptomatic Clustering in Adolescent Mental Health

**DOI:** 10.1101/2024.07.25.24311024

**Authors:** Michelle F. Kennedy, Paul Schwenn, Amanda Boyes, Lia Mills, Taliah Prince, Marcella Parker, Daniel F. Hermens

## Abstract

**Background:** Adolescence represents a sensitive developmental period characterised by an increased incidence of emerging mental health symptoms and formal diagnostic onset. These conditions can remain a significant burden throughout life. The Longitudinal Adolescent Brain Study (LABS) commenced in 2018 to track the onset and trajectory of mental health symptoms among general population participants. This research aims to identify polysymptomatic clusters of emerging mental health symptoms in adolescents and examine how these clusters vary by age and change over time, providing insights into the pluripotentiality of disorder development.

**Methods:** LABS participants aged 12-17 years (*n*=166) completed the Mini International Neuropsychiatric Interview (MINI Kid) approximately every 4 months, with up to 15 timepoints. Due to this high dimensional dataset, the data was first processed using a dimensionality reduction step (uniform manifold approximation and projection; UMAP). Following this, the data was clustered using Bayesian model averaging of k-means, gaussian mixture model and hierarchical clustering to identify distinct symptom clusters. Symptom clusters were described in terms of the original neuropsychiatric interview responses using separate XGBoost classifier models. Symptom cluster dynamics were analysed using Markov chain transition probability matrices and longitudinal analysis. To explore the relationship between symptom clusters and psychological distress and wellbeing, correlational analyses were conducted using scores from the Kessler Psychological Distress Scale (K10) and the COMPAS-W Wellbeing Scale.

**Outcomes:** Six symptom-based clusters (states) were identified: attention, anxiety, depression, manic episode - heritability, anhedonia, and well. Depression and anxiety clusters had the greatest pluripotentiality. Analysis of psychological distress and wellbeing demonstrated an inverse relationship between the states: those with greater psychological distress had more symptoms, conversely those with greater wellbeing had fewer symptoms.

**Interpretations:** Mapping polysymptomatic clusters of mental health symptoms and their pluripotential and transitory trajectories in adolescents enables more effective targeting of preventive interventions. This approach moves beyond categorical classifications to mitigate the progression of early symptoms into enduring psychiatric disorders.

## Introduction

Mental health disorders in adolescents represent a significant public health concern, contributing substantially to the global burden of disease^1^. The onset of many disorders commonly occurs during this critical developmental period with 34% by age 14 and 48% by age 18, significantly impacting individual well-being, academic performance, and social functioning^2^. While traditional diagnostic approaches have focused on categorical classifications, recent research emphasises the importance of considering transdiagnostic symptom dimensions and their developmental trajectories in adolescence^3,4^. A central tenet of the transdiagnostic concept is pluripotentiality, which recognises that early polysymptomatic presentations may lead to different manifestations of psychopathology over time^5^. This concept reflects the dynamic nature of symptom clusters and the potential for adolescents to transition between clusters during this developmental period^5^. It acknowledges both homotypic and heterotypic progression of polysymptomatic clusters, where early symptom presentation may lead to diverse outcomes^5^.

Transdiagnostic approaches to mental health have been proposed as a framework for incorporating the pluripotential and dimensional nature of mental illness, particularly important for youth populations where disorder onset often occurs^2,3^. These approaches examine the existence of transdiagnostic symptom clusters that transcend diagnostic categories, exploring common etiological pathways or risk factors^3^. By adopting this approach, researchers and clinicians can better understand the complex interplay of early symptoms across different disorders and potentially improve intervention strategies.

Longitudinal studies have provided valuable insights into the developmental trajectories of mental health disorders in adolescents^3,6^. However, further research is required to investigate the association between pluripotentiality, transdiagnostic polysymptomatic clusters and their longitudinal patterns. Utilising an agnostic approach that tracks emerging symptom clusters longitudinally can elucidate trajectories of adolescent mental health disorders^7^. Integrating key risk factors into polysymptomatic cluster modelling will support the selection of more targeted and effective treatment modalities individualised to each adolescent’s unique mental health presentation^7^. By adopting a dimensional and transdiagnostic perspective, researchers can explore the heterogeneity in symptom progression and the potential for diverse outcomes, reflecting the pluripotential nature of mental health disorders^8^.

This study employs a data driven approach, utilising complex statistical analyses, to identify patterns and explore the clustering of mental health symptoms in adolescents. The study’s primary objective is to identify polysymptomatic clustering of mental health symptoms in general population adolescents aged 12 – 17 years using a transdiagnostic and longitudinal framework. The identification of symptom clusters will enhance our understanding of emerging mental health symptoms and identify the pluripotential of clusters and individual trajectories of disorder development.

## Methods

The Longitudinal Adolescent Brain Study (LABS) was established to understand how the adolescent brain develops through ages 12-17 years and to investigate the trajectory of mental health symptoms.

### Participants

LABS uses longitudinal data collected from a community sample at the Thompson Institute, University of the Sunshine Coast (UniSC). Commencing in 2018, LABS collects participant data three times a year for up to five years, spanning ages 12 – 17 years. At each timepoint, data collected includes self-report questionnaires, cognitive testing, a neuropsychiatric interview, an Electroencephalogram (EEG), and Magnetic Resonance Imaging (MRI) (Supplementary material - figure S1). Demographic data collected includes date of birth, sex, gender, postcode, school, year, height, weight, mental health seeking/diagnoses/medications, major medical illnesses, hearing/vision and allergies. Participants primarily enter the study at 12 years of age, when they are in their first year of high school (year 7). In 2021, study entry criteria were expanded to 12-15 years at age of entry. LABS records sex via parental report at recruitment into the study (male/female/intersex), with the addition of self-reported sex and gender identity being added to the LABS protocol in March 2023.

Inclusion criteria were adolescents aged 12 years and in year 7 or up to 15 years of age, proficient in spoken and written English, able to attend onsite appointments at the Thompson Institute, and willing and able to complete the MRI. Exclusion criteria include adolescents who suffer from a major neurological disorder (such as epilepsy), intellectual disability, or medical illness, or who have sustained head injury (with loss of consciousness exceeding 30 minutes) as these conditions may compromise a participants’ safety and ability to undertake the LABS assessments. For a variety of reasons, (personal and family), some participants may not be able to attend every timepoint, in response, accommodations such as completing partial assessments or missing a timepoint are made. There were 176 participants in LABS at the time of analysis, of which 36 completed the five-year study, and 63 had withdrawn and 15 were lost to follow-up. Reasons for withdrawal included, parental work and participant school commitments, boredom with repeated protocol, participant individuation and changing priorities. Of the 176 participants, 166 fully completed the MINI Kid interview, with a total of *n* = 947 data points.

This observational cohort study was conducted in adherence with the Declaration of Helsinki and is approved by the UniSC Human Research Ethics Committee (A1811064). All participants and guardians provided informed written consent prior to commencement of study participation.

### Data Collection

With a focus on polysymptomatic clustering of emerging mental health symptoms and their pluripotential longitudinally, this study utilises LABS data collected from the Mini International Neuropsychiatric Interview (MINI Kid), the primary measure^9^, and the Kessler Psychological Distress Scale (K10) ^10^, and the COMPAS-W Wellbeing Scale ^11^, both secondary measures. Symptom level MINI Kid data was used to investigate polysymptomatic clustering transdiagnostically. The K10 was included to assess psychological distress, a well-established indicator of emerging psychopathology across various disorders^10^. The COMPAS-W was utilised to measure wellbeing, recognised as a potential protective factor against development of mental health disorders^11^. This dual focus on the variables psychological distress and wellbeing, reflects a growing emphasis on a more comprehensive, multidimensional approach to mental health diagnoses and treatment^12^. Self-report measures (K10 and COMPAS-W) are completed via a touch screen tablet, using the Qualtrics survey platform (Copyright © 2019 Qualtrics, Provo, UT, USA) in the presence of a research assistant (RA). Data included in this study was restricted to 166 unique participants from which MINI Kid raw interview question answers were available.

<u>MINI Kid</u> – is a structured neuropsychiatric interview assessing 30 major paediatric psychiatric disorders and comprises 417 items. RAs verbally administer the interview verbatim to the participant, providing clarification where necessary. Questions elicit specific diagnostic criteria, including symptom timeframe, frequency, and severity. The suicidality module was excluded from LABS due to the community sample (i.e., not a clinical population), and the inability to skip items based on previous responses, as in other modules, thereby significantly increasing the duration of the interview. Suicidality is assessed using a separate measure in the self-report questionnaire. Trained RAs administer the MINI Kid via the Proem Health digital platform, which incurs a per-interview fee. The digital format was selected to enhance data collection standardisation and coding efficiency given the extensive dataset^9^. The MINI Kid demonstrates high inter-rater and test-retest reliability (Kappa 0.64-1.00), and concurrent validity with other measures supporting its utility in assessing paediatric psychiatric disorders^13^.

<u>K10</u> – is a self-report measure of psychological distress consisting of 10 items assessing anxiety and depression symptoms in the last month. Answers are recorded on a 5-point Likert scale with responses ranging from 1 (none of the time) to 5 (most of the time). Total scores range from 1 -50 and are categorised as follows; Low (10-15), Moderate (16-21), High (22-29) and Very High (30-50). The K10 is brief, free of charge, and simple to administer. The K10 exhibits excellent internal consistency (Cronbach’s alpha >0.90) and good construct validity making it a reliable tool for screening psychological distress^10^.

<u>COMPAS-W</u> (secondary measure) – is a measure of wellbeing assessing poor to optimal wellbeing and resilience and comprises 26 items scored on a 5-point Likert Scale with responses ranging from 1 (strongly disagree) to 5 (strongly agree). A composite wellbeing score, the summation of all items, is categorised as follows; Languishing (≤ 88); Moderate (89-108) and Flourishing (≥ 109). The COMPAS-W is free of charge and simple to administer. The COMPAS-W scale demonstrates good internal consistency (Cronbach’s alpha =0.84) and good test-retest reliability (r=0.82), with strong convergent and discriminant validity supporting it as a reliable tool for measuring wellbeing^11^.

### Statistical Analysis

#### Dimensionality Reduction

Due to the high-dimensional nature of our data, we used Uniform Manifold Approximation and Projection (UMAP) ^14^ for dimensionality reduction, as it is known to preserve the global structure of the data (see Figure S2 for the processing pipeline flow chart).

Before UMAP embedding, MINI Kid interview responses were filtered and binarized to retain endorsements of specific symptoms (1 for "yes" and other text, 0 for "no," "unsure," and skipped questions with no symptoms in screener questions). Given the binary coding, a Hamming metric was used for the UMAP metric. UMAP embedding parameters were tuned by visually inspecting a 3D plot to assess the quality, topology, and spread of embeddings before clustering. Analysis was conducted using R (version 4.3.3, 2024-02-29).

This study exclusively focused on symptom-level question response data to avoid biasing the cluster algorithms with predefined diagnostic labels. For this reason, questions dependent on DSM diagnostic criteria were also removed. Of the 417 MINI Kid questions, 20 with constant responses across all participants were excluded before UMAP embedding. Questions deemed too general for most participants were dropped, including specific phobias (except a question about fears that cause problems). Questions concerning adjustment disorders were excluded due to missing data for the screener question inhibiting recoding of subsequent questions. Drug use questions were excluded due to low prevalence (3% responded). Body mass index (BMI) in Anorexia Nervosa was removed due to deemphasis of strict BMI cutoffs in the current classification system^15^. The Manic and Hypomanic Episodes (MHE) screener question was excluded due to misunderstanding and false positives. The MHE pre-screener question was included to investigate heritability’s influence on later psychopathology and mental health state transitions^8^. After the above exclusions, 274 MINI Kid questions remained which were then reduced to 3 UMAP embeddings.

#### Cluster Analysis

##### Model Averaging Clustering

From the UMAP embeddings, the R package *ClusterBMA*^16^ was used to determine robust cluster assignment from k-means, hierarchical, and Gaussian mixture model clustering methods. The optimal number of clusters was determined based on two criteria: (1) the highest silhouette score and (2) the ability to produce a "well" cluster with maximum proportion of ‘no’ responses indicating a predominant absence of symptoms – a known existing condition in the population. Herein, a cluster is interpreted as a distinct mental health “state” (total *k* states). A participant can only be allocated to a single state at a given timepoint but can transition between states across timepoints.

##### State Characterization/Labelling

Once the optimal number of states were identified, state features were extracted by fitting an XGBoost (extreme gradient boosting) classifier to each cluster. XGBoost is a machine learning ensemble method that is well-suited to high-dimensional data due to L1 and L2 regularisation (penalises the number of branch weights and large weight values) which can be tuned to reduce overfitting on data with multicollinearity^17^. The binary logistic classifier target was defined by setting those in the given state to 1 and those outside the state to 0 and coded responses as predictors. The importance of each feature (question/symptom) can be understood in terms of the gain metric of XGBoost (validated using leave one group (ID) out cross validation). By considering both the feature importance and proportion of affirmative answers in each cluster, we could determine which symptoms characterise each state based on their presence or absence.

##### Symptom Diversity Index

The average number of “yes” responses within each state defined the symptom diversity allowing us to identify the most ‘polysymptomatic’ state. A symptom diversity index was formed by dividing by the state with largest symptom diversity. A value of zero means the state is characterised by no symptoms.

##### Pluripotential Index and state transitions

States were defined globally, and a state transition probability matrix was calculated using Markov chain modelling. We introduce a pluripotential index from the Shannon entropy of each row of the transition probability matrix. The maximum pluripotential occurs when there is equal probability of transitioning to any state in the matrix (see Equation S1). Transition probabilities in the Markov matrix were categorized as low (0.00-0.33), moderate (0.34-0.66), or high (0.67-1.0) to facilitate interpretation of state transition likelihoods. The stationary distribution was calculated as the left eigenvector corresponding to the eigenvalue of 1 for the transition matrix.

##### Psychometric Assessment

Once states were labelled and features identified, the *survey* package in R^18^ was used to estimate the means and confidence intervals for K10 score and COMPAS-W Wellbeing score.

## Results

There was a total of 166 unique participants in the study: 93 females (56%) and 73 males (44%), 0 other, with 947 total observations longitudinally. The breakdown longitudinally by number of participants is displayed in Table 1 and Figure 1. Of the 166 participants, 57.8% were mental health help-seeking, with a higher proportion of female participants (34.9%), compared to males (22.9%) (Tables S2-S5)

**Figure 1:**
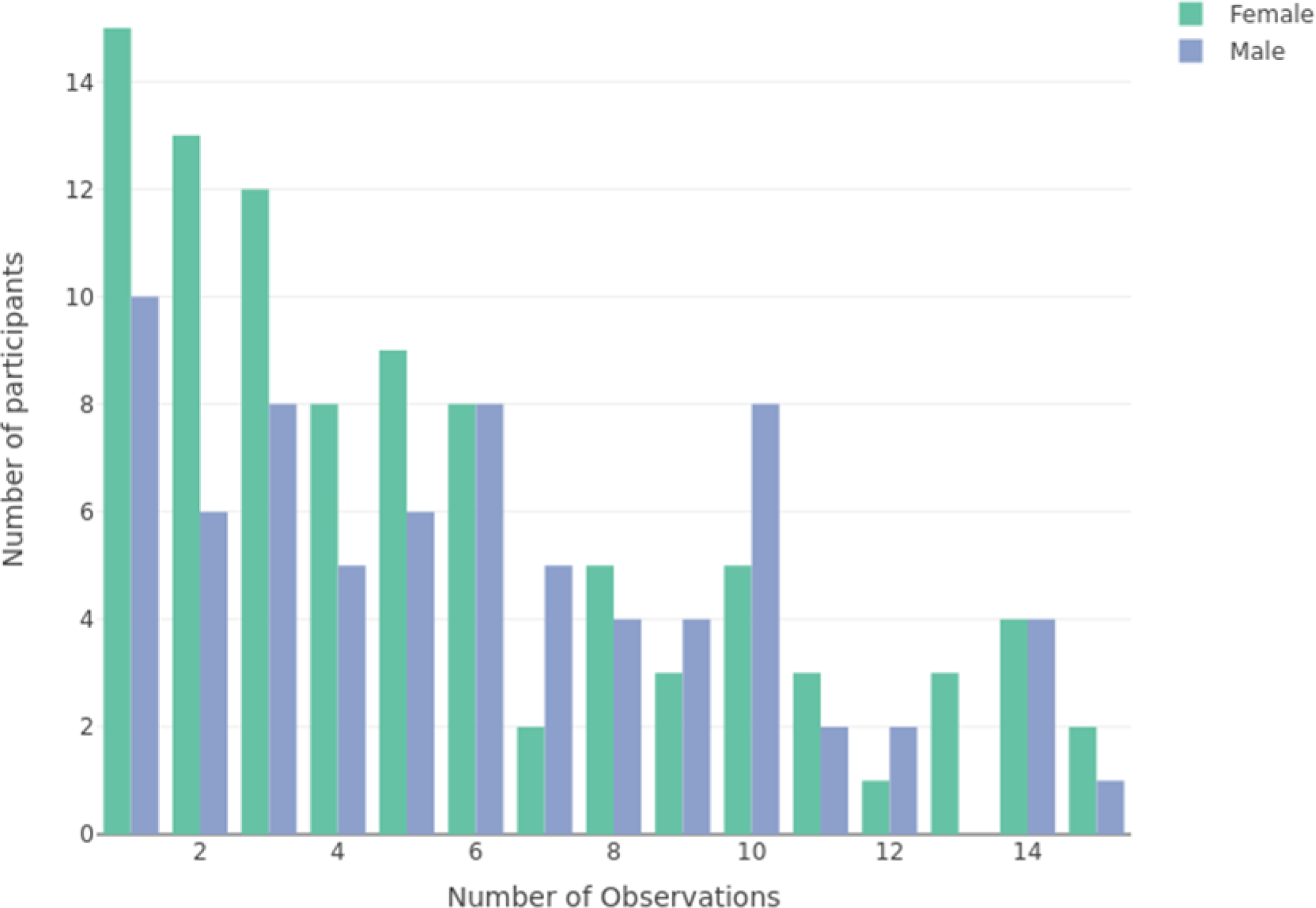
Number of observations by number of participants and sex.

**Table 1:** Number of observations as a function of number of participants.

| **Number of observations (timepoints)** | 1 | 2 | 3 | 4 | 5 | 6 | 7 | 8 | 9 | 10 | 11 | 12 | 13 | 14 | 15 |
| --- | --- | --- | --- | --- | --- | --- | --- | --- | --- | --- | --- | --- | --- | --- | --- |
| **Total number of participants** | 25 | 19 | 20 | 13 | 15 | 16 | 7 | 9 | 7 | 13 | 5 | 3 | 3 | 8 | 3 |
| **Females** | 15 | 13 | 12 | 8 | 9 | 8 | 2 | 5 | 3 | 5 | 3 | 1 | 3 | 4 | 2 |

Interpretation of states using our longitudinal data requires clarification of the following *N* refers to the total number of unique participants that have occupied a state (cluster) at any time longitudinally. States identified were (1) Well, (2) Depression, (3) Anxiety, (4) Attention, (5) Manic Heritability, and (6) Anhedonia (Table 2 and Figure S4).

**Table 2:** Key symptoms defining each state (highest feature importance for cluster assignment)

| Well (1) | Depression (2) | Anxiety (3) | Attention (4) | Manic Heritability (5) | Anhedonia (6) |
| --- | --- | --- | --- | --- | --- |
| Absence of psychopathology | Exclusively depression | Excessive worry | Difficulties in behaviour, restlessness and inattentiveness | Family history of: manic-depressive bipolar disorder | Diminished capacity to experience pleasure |
| No depression | Persistent low mood | Anxious feelings | No depression | Mood swings treated with medication | No sadness, irritability or hopelessness |
| Sub-emerging feelings of anxious, scared or uneasy in places/situations where might become frightened | Irritability | Sensory processing sensitivity |  | No depression |  |
|  | Tiredness | No depression |  |  |  |
|  | Past episodes of depression |  |  |  |  |

Of the 166 participants, 62 were in one state, 51 transitioned between two states, 43 between three states, 9 between four states, and 1 transitioned between five different states over various number of observations.

### State descriptives

Figure 5 shows a 3D projection of the three UMAP embeddings. The Bayesian model averaged state solution found good separation for all six of states with larger markers indicating greater model allocation uncertainty. Each state was clearly identified from the predominant XGBoost feature importance gain (see Figure S4).

### State proportions

Table 3 describes state stationary distribution (long term occupancy probability), state prevalence (participants visiting the state at any time). We will use the notation *x*% (*f,m*) where x is the percent of total participants that have occupied the state at any time, *f* is the % of total females that have occupied the state at any time and similarly *m* for males. State occupation proportions are further defined below.

**Table 3:**
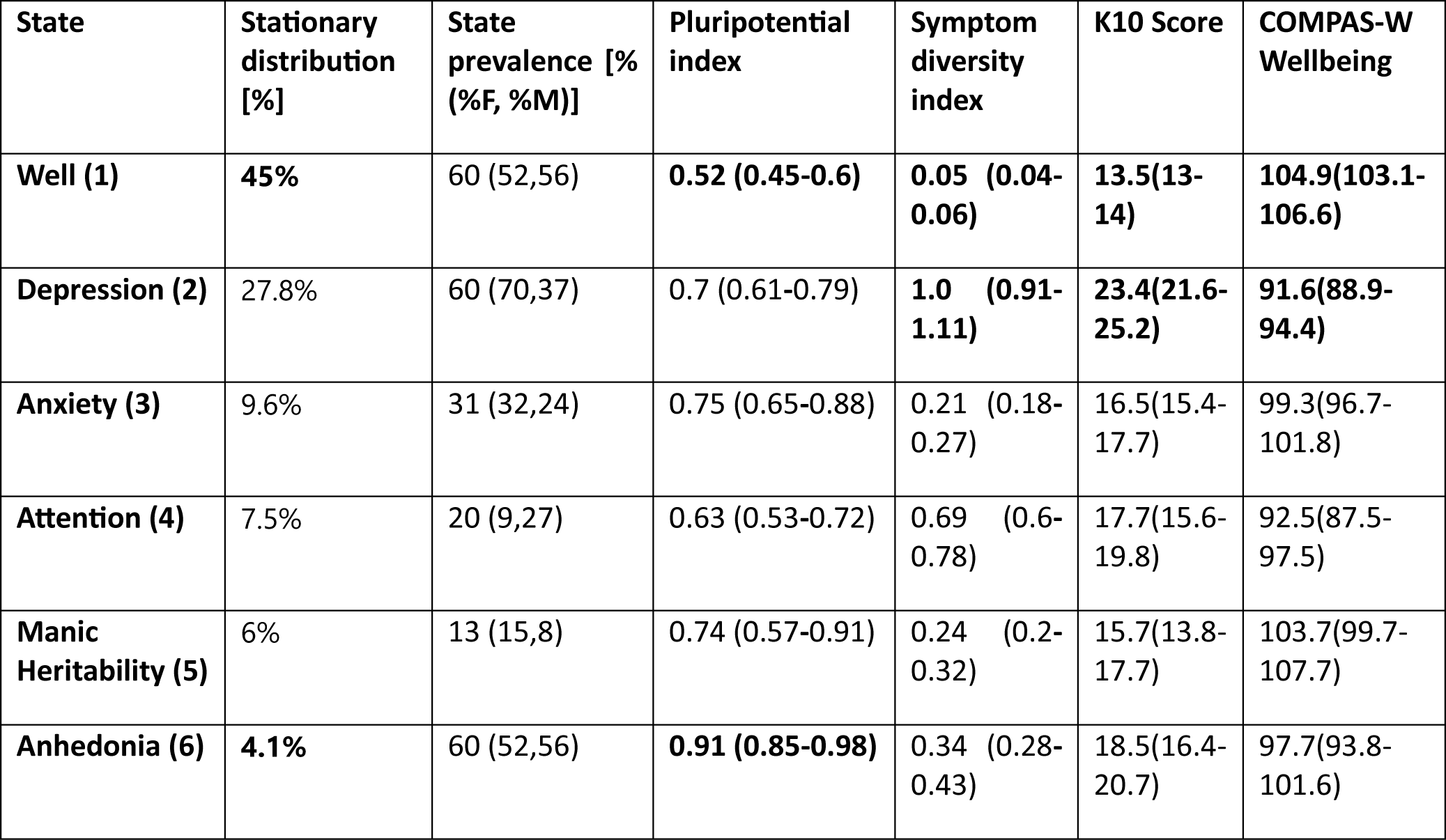
State descriptives. Stationary distribution (long term occupancy probability), state prevalence (participants visiting the state at any time), pluripotential index (state transition uncertainty), symptom diversity index, average k10 score and COMPAS-W Wellbeing. 95% CIs in brackets. Bold numbers represent the lowest and largest values within state descriptives.

| State | Stationary distribution [%] | State prevalence [% (%F, %M)] | Pluripotential index | Symptom diversity index | K10 Score | COMPAS-W Wellbeing |
| --- | --- | --- | --- | --- | --- | --- |
| **Well (1)** | **45%** | 60 (52,56) | **0.52 (0.45-0.6)** | **0.05 (0.04-0.06)** | **13.5(13-14)** | **104.9(103.1-106.6)** |
| **Depression (2)** | 27.8% | 60 (70,37) | 0.7 (0.61-0.79) | **1.0 (0.91-1.11)** | **23.4(21.6-25.2)** | **91.6(88.9-94.4)** |
| **Anxiety (3)** | 9.6% | 31 (32,24) | 0.75 (0.65-0.88) | 0.21 (0.18-0.27) | 16.5(15.4-17.7) | 99.3(96.7-101.8) |
| **Attention (4)** | 7.5% | 20 (9,27) | 0.63 (0.53-0.72) | 0.69 (0.6-0.78) | 17.7(15.6-19.8) | 92.5(87.5-97.5) |
| **Manic Heritability (5)** | 6% | 13 (15,8) | 0.74 (0.57-0.91) | 0.24 (0.2-0.32) | 15.7(13.8-17.7) | 103.7(99.7-107.7) |
| **Anhedonia (6)** | **4.1%** | 60 (52,56) | **0.91 (0.85-0.98)** | 0.34 (0.28-0.43) | 18.5(16.4-20.7) | 97.7(93.8-101.6) |

The ‘Well’ state with 60% occupancy (*f*52, *m*56), was defined by the absence of major psychiatric symptoms, specifically complete absence of depression screener response (question A1a) and absence of any depressive episodes (A6). ‘Depression’ with 60% occupancy (*f*52, *m*56), was characterised by a strong proportion of responses to the depression screener in conjunction with depressive episodes. ‘Anxiety’, with 31% occupancy (*f*32, *m*24), was defined by a strong response to the agoraphobia screener followed by a low response to autism symptoms (≈11% question X5) and the depression screener (≈ 8% question A1a), with no attention or depression symptoms evidenced. ‘Attention’ with 20% occupancy (f9, m27), was characterised by a strong proportion of responses to the attention screener and complete absence of the depression screener response (question A1a). ‘Manic Heritability’, with 13% occupancy (15, 8), was defined by a strong response to manic heritability (question C-unscored), and complete absence of depression screener response (question A1a). Finally, ‘Anhedonia’, with 60% occupancy (f52, m56), was characterised by a strong proportion of responses to the anhedonia depressive symptom (question A2a), in contrast to a complete absence of depression screener response (question A1a).

### Pluripotential and Symptom Diversity

**Pluripotential index** refers to the transition probability uncertainty between states^19^. The greatest pluripotential (0.9) was found in the Anhedonia state with the lowest pluripotential found for the Well state (0.5) (Table 3 & Figure 2).

**Figure 2:**
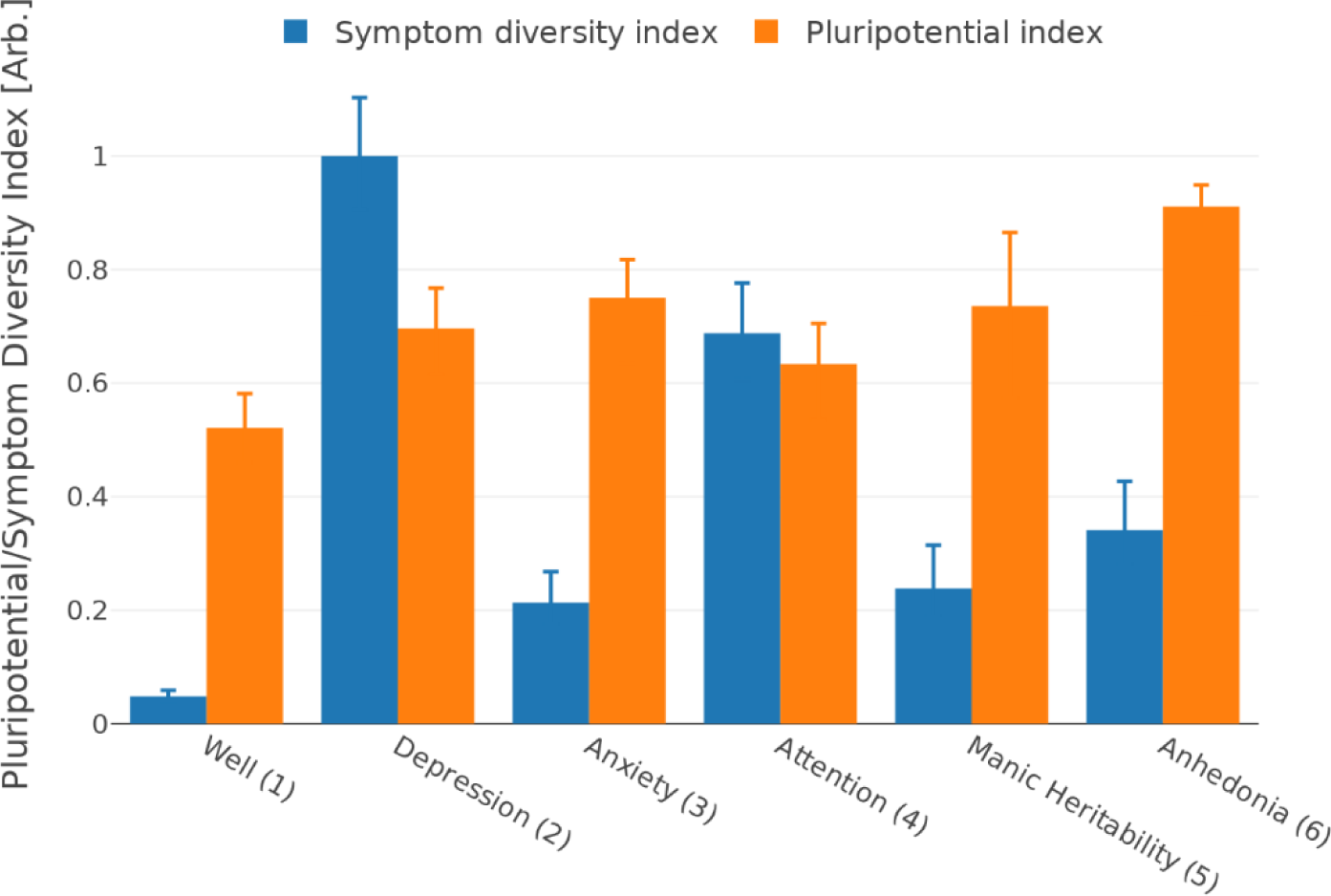
Pluripotential index and symptom diversity index by state.

**Symptom diversity** refers to the amount of transdiagnostic symptoms within a state. Greater symptom diversity was associated with greater psychological distress and lower wellbeing, evidenced in the Depression state, and conversely for lower symptom diversity, observed in the Well state. Thus, symptom diversity is a strong indicator of wellbeing and psychological distress (Table 3, Figure 2). Figure 6, panel A, shows that states experiencing higher psychological distress exhibited more symptoms (larger markers, r^2^ = 0.83, p < 0.02), and states with greater wellbeing showed fewer symptoms (smaller markers, r^2^ = 0.85, p < 0.01). Panels B and C detail psychological distress (wellbeing) vs symptom diversity separately for females r^2^ = 0.82, p < 0.02 (r^2^ = 0.75, p < 0.03) and males, r^2^ = 0.78, p < 0.02 (r^2^ = 0.96, p < 0.001).

### State Proportion by sex and age

Table 4 shows the total count of unique participants in each age group and sex. Most participants were found in the Well and Depression states, with equal representation from male and female sexes in the Well state. However, females over-represent in Depression, Anxiety, Manic Heritability and Anhedonia states, whilst males over-represent in the Attention state. The smallest representation was Manic Heritability state.

**Table 4:** Unique participants in each state by age group [total (females, males)]

| Age group | Well (1) | Depression (2) | Anxiety (3) | Attention (4) | Manic Heritability (5) | Anhedonia (6) |
| --- | --- | --- | --- | --- | --- | --- |
| 12 | 47 (25,22) | 26 (19,7) | 18 (11,7) | 5 (0,5) | 2 (2,0) | 7 (4,3) |
| 13 | 65 (32,33) | 32 (23,9) | 27 (15,12) | 18 (4,14) | 12 (9,3) | 11 (6,5) |
| 14 | 51 (24,27) | 41 (28,13) | 15 (6,9) | 15 (4,11) | 8 (5,3) | 5 (4,1) |
| 15 | 38 (18,20) | 35 (23,12) | 7 (3,4) | 10 (1,9) | 8 (5,3) | 6 (4,2) |
| 16 | 22 (9,13) | 31 (19,12) | 4 (3,1) | 2 (1,1) | 7 (4,3) | 4 (2,2) |
| 17 | 14 (8,6) | 17 (9,8) | 6 (4,2) | 2 (0,2) | 6 (4,2) | NA |

Figure 3 plots the proportion of each state by age group. The proportion assigned in the Well state *decreases* with age group whereas the proportion assigned in the Depression state *increases* with age group. The proportion assigned in the Anxiety state decreases with age, however a slight increase occurs at age 16 years. The proportions assigned to the Attention, Manic Heritability & Anhedonia states remain relatively constant across age groups, with a slight increase at age 15 years for the Manic Heritability state and a slight decrease at age 15 years for the Attention state.

**Figure 3:**
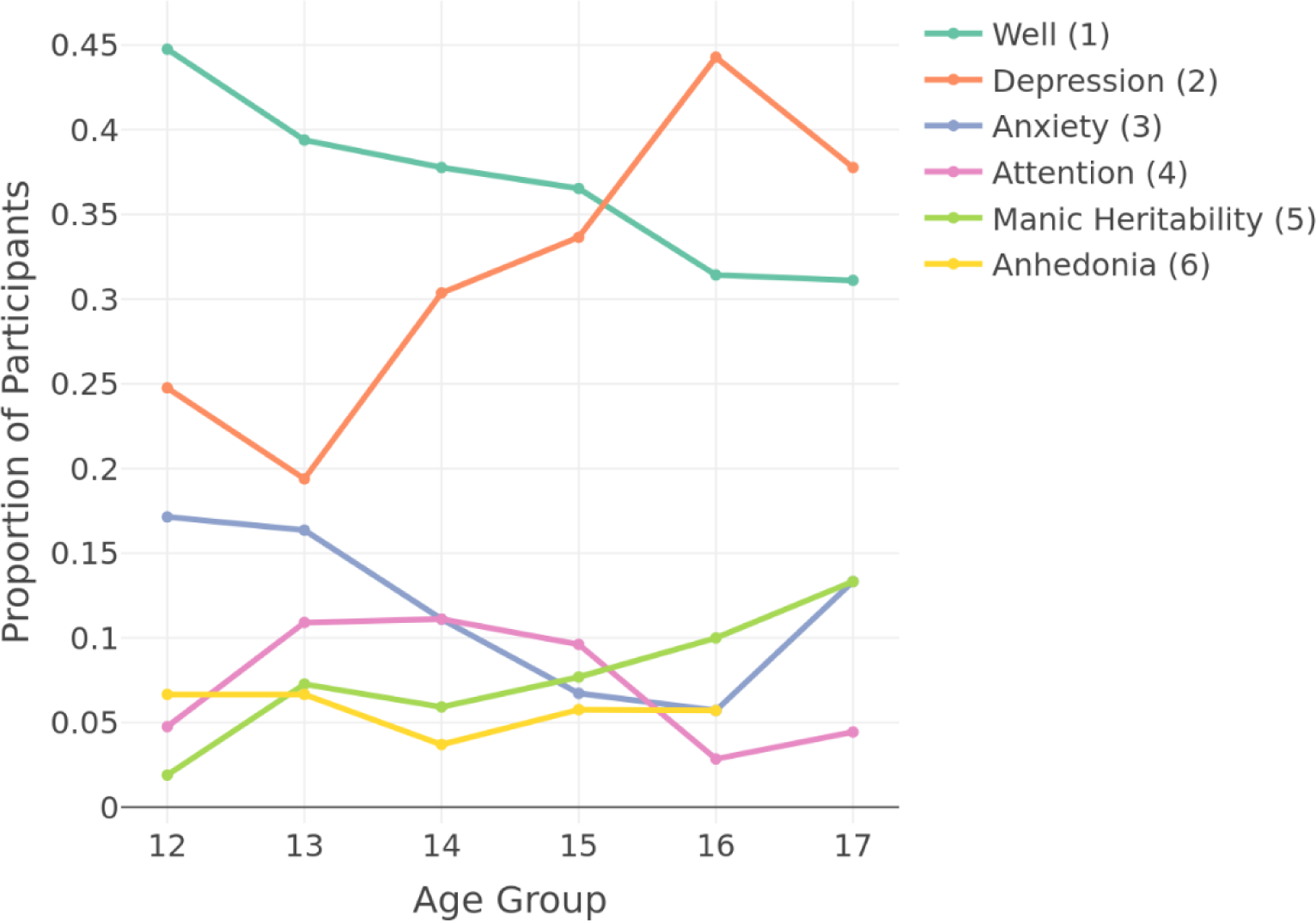
Proportion of participants (%) in each state by age group.

### State transition probabilities

The change in state allocation or proportions in the transition probability matrix (Figure 4), can be understood in terms of trajectories between states. The decrease in Well proportion is due to predominant transitions from Well to Depression [probability 0.11(95% CI 0.08-0.14)] and to Anxiety [0.08 (95% CI 0.05-0.11)] with probability of remaining Well high [at 0.74 (95% CI 0.65-0.83)]. Depression had low to moderate transitions from Attention [0.26 (95% CI 0.13-0.40)], Manic Heritability [0.26 (95% CI 0.11-0.4)] and Anhedonia [0.25 (95% CI 0.06-0.44)] with a lower probability of transitioning to Well [0.15 (95% CI 0.10-0.21)] and moderate to high probability of remaining in Depression [0.61 (95% CI 0.51-0.72)]. The probability of remaining in Anxiety was moderate [0.29 (95% CI 0.18-0.41)], with the highest transition from Anhedonia [0.11 (95% CI 0.00-0.23)] with a moderate probability of transitioning to Well [0.46 (95% CI 0.32-0.61)]. Likewise, Attention had the highest transition probability from Anhedonia [0.11 (95% CI 0.00-0.23)], with highest probability of transitioning to Depression [0.26 (95% CI 0.13-0.40)’ and Well [0.16 (95% CI 0.05-0.26)], however there is a higher probability of remaining in Attention [0.53 (95% CI 0.34-0.71)]. Similarly, the probability of remaining in Manic Heritability is moderate [0.50 (95% CI 0.30-0.70)], with the highest transition from Depression [0.06 (95% CI 0.03-0.70)] and highest probability of transitioning to [Depression 0.26 (95% CI 0.11-0.41)] followed by Well [0.13 (95% CI 0.03-0.23)]. Anhedonia has similar moderate probabilities of transitions to Depression [0.25 (95% CI 0.06-0.44)] and Well [0.29 (95% CI 0.09-0.48)], with the probability to remain in Anhedonia being low [0.21 (95% CI 0.04-0.39)].

**Figure 4:**
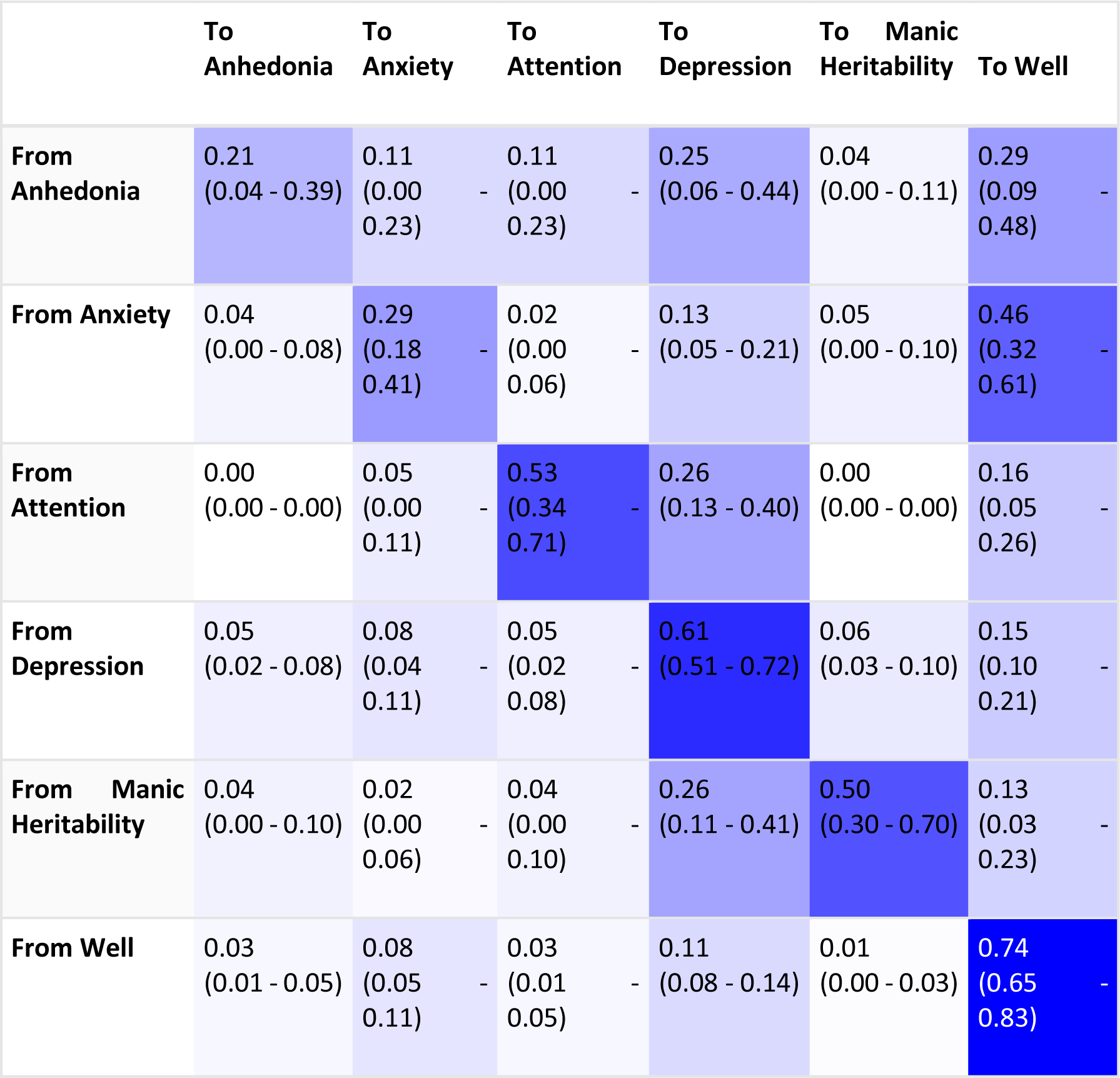
Heat map of state transition probabilities between states (95% CI’s in brackets), at any stage of the longitudinal study (up to 15 timepoints). Darker elements represent greater probability of transition from a given state (rows) to any other state (columns).

### States, psychological distress and wellbeing

Figure 6 illustrates the relationship between wellbeing and psychological distress across different states. Panel A shows an inverse relationship between state wellbeing and distress (r^2^ = 0.72, p < 0.04). Panels B and C detail these relationships separately for females (r^2^ = 0.73, p < 0.03) and males (r^2^ = 0.84, p < 0.01). The symptom states were analysed in relation to psychological distress and wellbeing, revealing heterogenous outcomes (means) that varied across sex and age groups (Figure 5 and 10, Table S8). For psychological distress, distinct patterns emerged whereby the Well state showed a gradual mean increase with age, while the remaining states had various peak ages: Depression (peaks at ages 13, 16 & 17), Anxiety (peaks ages 12 & 15), Attention (peaks ages 14 & 16), and Manic Heritability (peaks 13 & 15) while Anhedonia had a single peak at age 14. Females had a higher probability of psychological distress across states. For wellbeing, the Well state exhibited high moderate wellbeing across all ages, whilst the Depression state displayed low moderate wellbeing across all ages, with borderline levels from 13-17 years. Anxiety and Manic Heritability states exhibited moderate wellbeing levels across all ages. The Anhedonia state displayed low moderate wellbeing levels across all ages. The Attention state demonstrated moderate wellbeing at ages 12,13 & 15, but languished at ages 14, 16 & 17. Minimal sex differences were observed in wellbeing, with males exhibiting a slightly higher probability in the anxiety state. An inverse relationship was also found between state, psychological distress and wellbeing being higher psychological distress had more symptoms and higher wellbeing had less symptoms (Figure 3). The Depression state had high psychological distress and low moderate wellbeing, with the Attention state having moderate psychological distress and low moderate wellbeing. Anhedonia and Anxiety states had moderate psychological distress and moderate wellbeing. Manic Heritability state had moderate psychological distress and high moderate wellbeing, with the Well state displaying low psychological distress and high moderate wellbeing.

**Figure 5:**
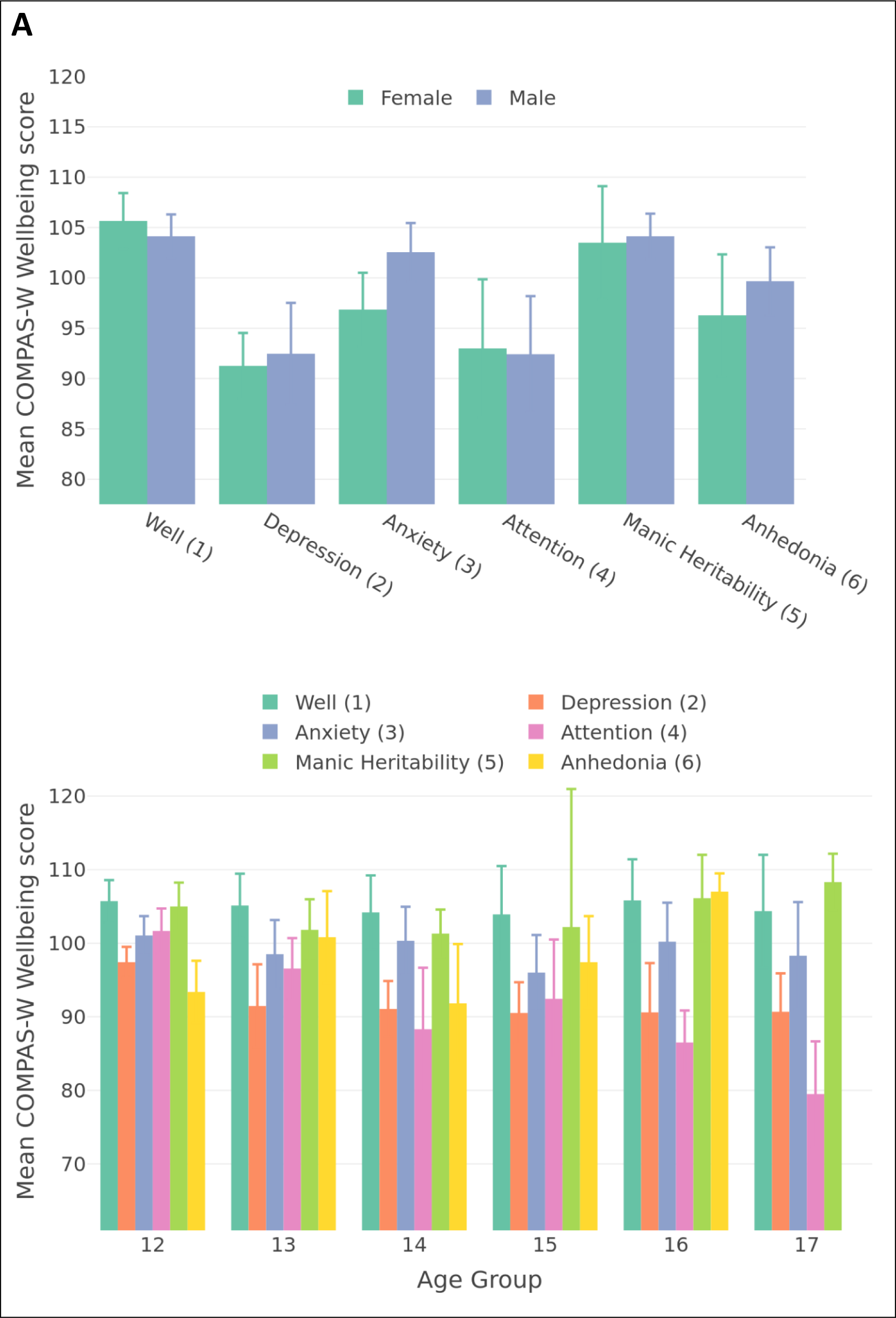
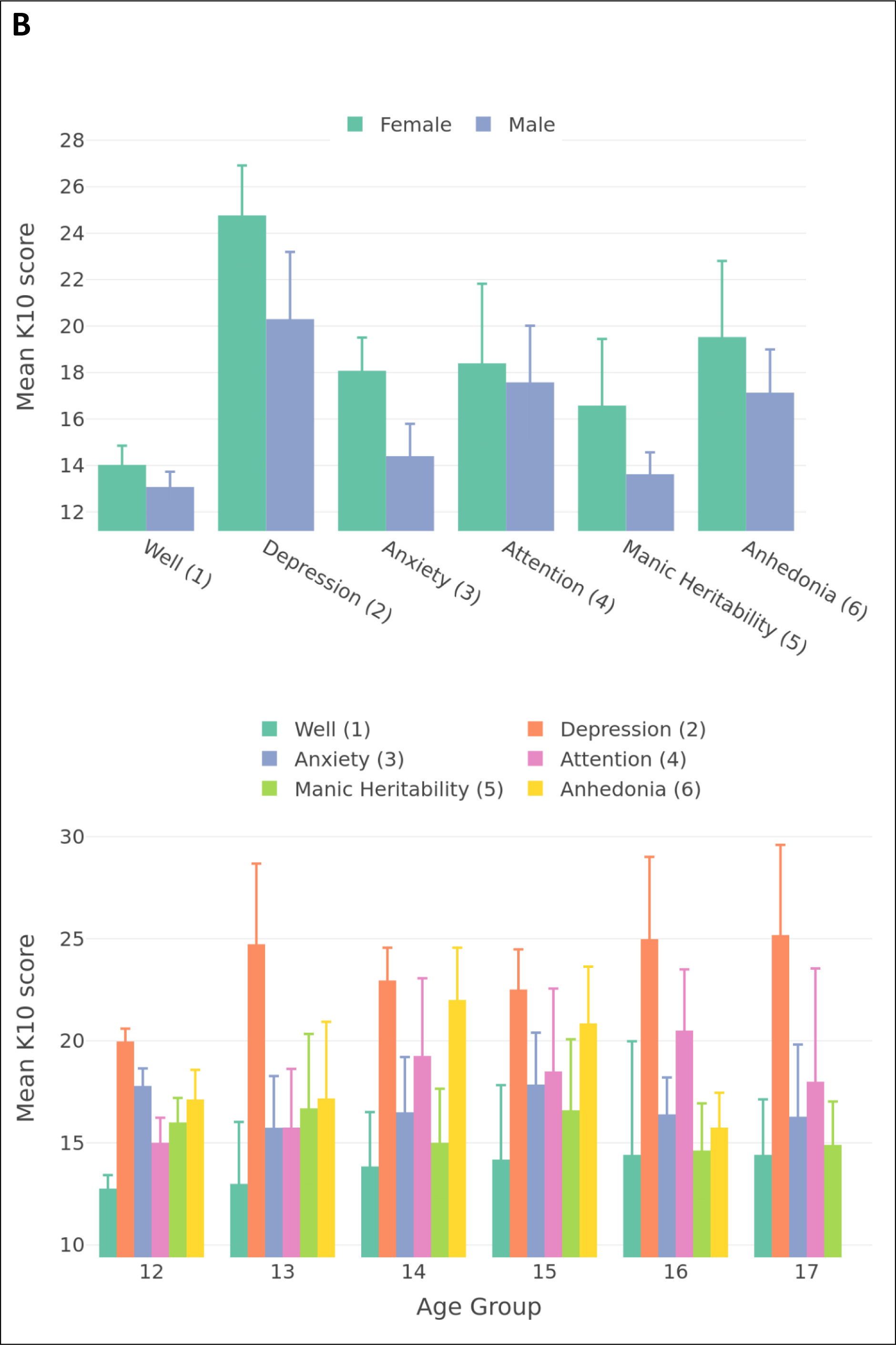
State means by age group (bottom) and sex (top) for K10 score (A) and COMPAS-W (B)

**Figure 6:**
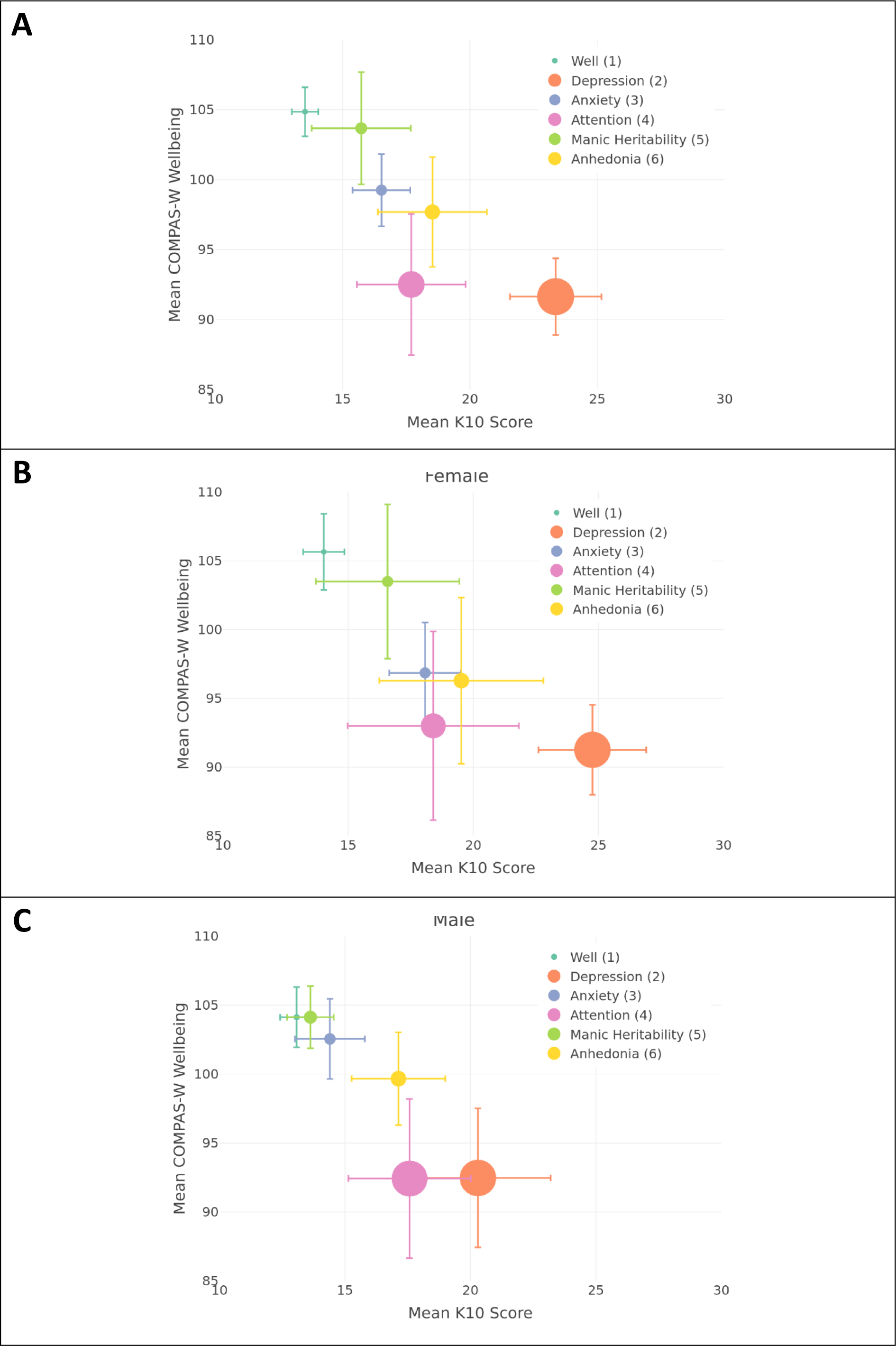
(**A**) Mean COMPAS-W Wellbeing score vs Mean K10 Score (psychological distress) by state. There was an inverse relationship between state, psychological distress, and wellbeing. In addition, states with greater psychological distress had more symptoms (larger marker size). Conversely states with greater wellbeing had fewer symptoms (smaller marker size). (**B**) Female states, and (**C**) Males states. Error bars denote 95% CIs.

## Discussion

Our data-driven analyses, utilising LABS data, identified six distinct symptom-based clusters, conceptualised as states: Anxiety, Anhedonia, Attention, Depression, Manic Episode - Heritability, and Well. These states align with the dimensional frameworks Hierarchical Taxonomy of Psychopathology (HiTOP) spectra and the Research Domain Criteria (RDoC) biobehavioural dimensions, reflecting the models’ emphasis on dimensional psychopathology and transdiagnotic phenotypes^3,4^. Specifically, the states align with the HiTOP Internalising spectrum (anxiety, depression and anhedonia) and the Disinhibited Externalising spectrum (attention), and RDoC domains of Negative valence systems (anxiety, depression and anhedonia) and Cogntive systems (attention) ^3,4^. The Manic Episode – heritability state whilst not listed as a symptom under these frameworks has been suggested as a biological symptom as genetic factors contribute to various psychopathologies^4^. Additionally, analyses revealed pluripotentiality across mental health states, highlighting the fluidity of transitions between states. The Well state was characterized by high self-transition probability such that once an individual transitioned into this state, they had a high probability of remaining well. In addition, the clustering captured variability of symptom diversity across states, and the state symptom diversity was associated with varying levels of psychological distress and wellbeing. The Well state had the highest long term occupancy probability and the Anhedonia state had the least. State transitions quantified pluripotentiality, with Anhedonia being associated with the highest level and the Well state with the lowest.

The Well state, demonstrated a high prevalence with balanced sex distribution^11^. The proportion assigned to this state decreased from age 14, aligning with evidence of increased mental health issue prevalence after this age^2^. The Well state exhibited the lowest pluripotentiality and symptom diversity, and largest stationary distribution demonstrating a tendency for maintenance in this state^7^. This suggests that early targeted interventions could potentially mitigate the risk of transitioning to alternative psychopathological states and faciliate maintenance in well^7^. The Well state is characterised by the inverse relationship of low psychological distress and high moderate wellbeing scores, which substantiates the importance of targeted interventions to reduce psychopathology and enhance positive mental health in adolescence^12^.

The Depression state exhibited a high prevalence, with a higher occurrence in females aligning with well established research on the sex disparity, particularly after age 12. The proportion assigned increased with age, particularly in females which is consistent with established trajectories in adolescence^20^. Transition patterns showed highest probability to well, suggesting potential recovery, while there were equal transitions from anhedonia, attention and manic heritability. Depression demonstrated moderate pluripotentiality and highest symptom diversity, reflecting the heterogeneous nature of symptom presentations, suggesting a heightened risk for comorbidity and transition to this state^21^. Characterised by high psychological distress and one of the lowest wellbeing, this state presents significant risk factors for symptom maintenance, particularly in females. Research suggests this may be due to the complex interplay of biological, psychological and social factors, specifically pubertal hormonal changes and heightened stress reactivity commonly experienced by female adolescents^22^.

The Anhedonia state demonstrated a high prevalence with a balanced sex distribution and remained constant across age groups in contrast to typical depression patterns. This suggests anhedonia may have distinct etiological pathways^23^. Transition patterns revealed highest probabilities to Depression and Well states, suggesting a pivotal role for the Anhedonia state in psychopathological trajectories and recovery^24^. Equally low transition probabilities from other states, suggest Anhedonia as a state reflecting a distinct psychopathological process rather than a common endpoint of other symptom progressions^24^. Anhedonia exhibited low symptom diversity and the highest pluripotentiality consistent with the lowest stationary distribution, reflecting its transdiagnostic nature as a common symptom across a variety of psychological disorders including depression, schizophrenia, anorexia nervosa and substance use disorders as well as anxiety and attention-deficit/hyperactivity.^24^. Characterised by moderate psychological distress and low moderate wellbeing, the Anhedonia state may represent a risk for transitioning to other states.

The Anxiety state had moderate prevalence, supporting well established research of higher prevalence in females commonly associated with increased anxiety sensitivity, higher reactivity to stress and pubertal changes^20^. The proportion assigned decreased with age, with an increase at age 16, reflecting complex developmental trajectories during this dynamic period^20^. Transition patterns revealed highest probability of moving to Well, then Depression, aligning with probable anxiety remission and progression (higher for females) in adolescents^20^. Low transitions from other states may indicate its role as a transdiagnostic state with diverse developmental trajectories in adolescent psychopathology^25^. Of note, Anxiety demonstrated moderately high pluripotentiality with low symptom diversity, supporting the transdiagnostic nature and core feature across different states^25^. The Anxiety state is characterised by moderate psychological distress and low moderate wellbeing which suggests ‘languishing’in the dual continuum model^12^. Thus interventions should target both symptom reduction and wellbeing.

The Attention state demonstrated a low prevalence, with a higher prevalence among adolescent males^26^. The proportion remained relatively stable across age groups, showing a slight decrease at age 15, indicating the persistence of attention-related symptoms throughout adolescence^27^. Transition patterns revealed highest probability to Depression, particularly in males, aligning with research on ADHD-depression comorbidity^28^. Furthermore, this transition risk is characterised by moderate psychological distress and wellbeing, highlighting the complex interplay between attention difficulties and depressive symptoms in adolescents^28^. The low transition probabilities from other states, suggest that attention-related difficulties represent may a distinct psychopathological process rather than a progression from other states. The Attention state exhibited moderate pluripotentiality and symptom diversity, indicating that while it has a set of core symptoms, it can manifest with a variety of other symptoms, highlighting the state’s transdiagnostic nature^28^.

The Manic episode-heritability state showed low prevalence, with a higher occurrence of females, consitent with bipolar spectrum disorders^29^. The proportion assigned remained relatively constant across age groups, increasing slightly from age 15, reflecting the emergence of manic/bipolar symptoms in late adolescence^29^. Notably, this state lacked depressive symptoms, which may be associated with the non-specific and transdiagnostic early symptom presentation^29^. However, transition patterns revealed highest probability to Depression, then Well states, and low transition probabilities from other states support the notion of strong genetic underpinnings^29^. Characterised by moderate psychological distress and wellbeing, personalised interventions should address early bipolar symptoms (e.g. cyclothymic) and wellbeing, due to the increased heritability risk for this state^29^.

The findings across various states highlight the complex and dynamic nature of early symptomatology of adolescent mental health. The Well state emerged as a crucial protective factor, emphasizing the importance of maintaining and promoting wellbeing. Depression and Anxiety states demonstrated sex-specific vulnerabilities, highlighting the need for sex-sensitive interventions, with anxiety considered a transdiagnostic symptom. Anhedonia had the highest pluripotentiality and appeared as a potential early indicator and transdiagnostic symptom, whilst the Attention state showed persistence and potential for comorbidity development. The Manic episode-heritability state revealed evidence in support of increased heritability risk, particularly in females. Collectively these findings emphasise the symptom diversity and the pluripotential for state transitions among adolescents and the importance of early identification, with personalised interventions that consider sex differences. The promotion of wellbeing emerged as a common theme across states due to its protective factor for addressing emerging adolescent mental health symptoms.

Several limitations warrant consideration. First, the protracted nature of longitudinal research presents challenges in maintaining participant retention and attrition bias: barriers to continuation have been addressed where possible for example by reducing frequency of timepoints, inviting participants to return for final timepoints, and providing travel reimbursement where travel costs were the primary barrier. Second, the study population may exhibit selection bias due to the potential overrepresentation of mental-health help-seeking participants, which could overestimate the prevalence or severity of mental health symptoms in adolescent populations. However, it should be noted that LABS researchers do not provide interventions, but refer participants to appropriate services when additional support is deemed necessary. Third, to mitigate interviewer-induced bias and enhance inter-rater reliability during administration of the MINI-Kid, RA’s underwent training and participated in regular discussion sessions. This protocol aimed to ensure that observed longitudinal changes in participants’ mental health states reflected symptom fluctuations rather than variability in interview administration or interpretation. Next, generalisability of findings may be limited due to small sample size and unaccounted cultural and socioeconomic factors may indicate a potential selection bias. Finally, conducting the interview 4 monthly over 5 years may introduce practice effects through participant familarisation of the tool, potentially leading to response bias and spurious symptom reduction.

Future research should incorporate larger-scale longitudinal studies with neuroimaging and genetic analyses to elucidate the neurobiological underpinnings of transdiagnostic symptom clusters/states across adolescent developmental trajectories. This will inform development of personalised interventions to maintain well states and prevent the progression to severe psychopathological states.

In conclusion, our findings add to the emerging research supporting a dimensional and transdiagnostic approach to mental health symptoms by providing a more dynamic understanding of the emergence and progression of adolescent psychopathology. The identification of and transitions between states of varying levels of pluripotentiality and symptom diversity align with conceptualisations of adolescent psychopathology as complex, interconnected symptom networks rather than discrete diagnostic categories.

## Author contributions

Michelle F. Kennedy: methodology, investigation, formal analysis, writing –original draft, review and editing. Paul Schwenn: statistical methodology, formal analysis, writing – review and editing. Amanda Boyes: data collection, writing – review and editing. Lia Mills: data collection, writing – review and editing. Taliah Prince – data collection, writing – review and editing. Marcella Parker: data collection, data curation, writing – review and editing. Daniel Hermens: conceptualisation, data curation, formal analysis, writing – review and editing, and supervision.

## Acknowledgements

The authors would like to thank the LABS study participants and their caregivers who made this work possible.

## Author Approval

all authors have seen and approved the manuscript.

## Data Availability

the data and code necessary to reproduce the analyses here are not publicly accessible, as are materials necessary to replicate the findings. Analyses were not preregistered.

