## Supplementary material for "Longitudinal Dynamics and Pluripotentiality of Polysymptomatic Clustering in Adolescent Mental Health"

#### Table of Contents


1. Supplementary Methods
2. Supplementary Results


### 1. Supplementary Methods

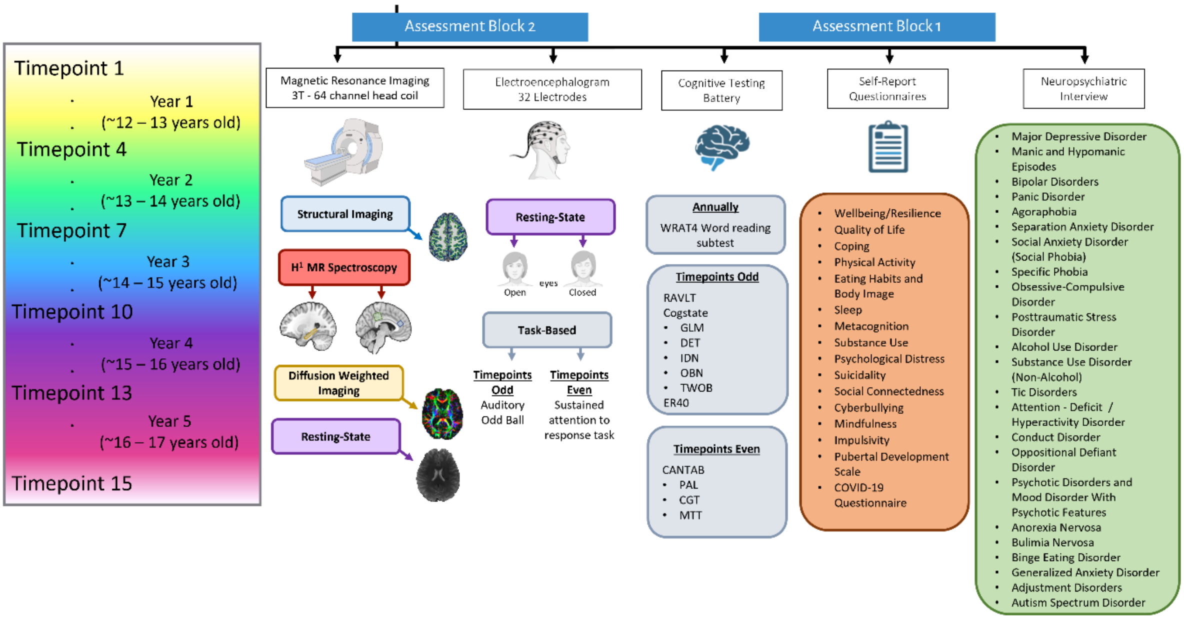


**Figure S1 Overview of Data collected at each timepoint.**

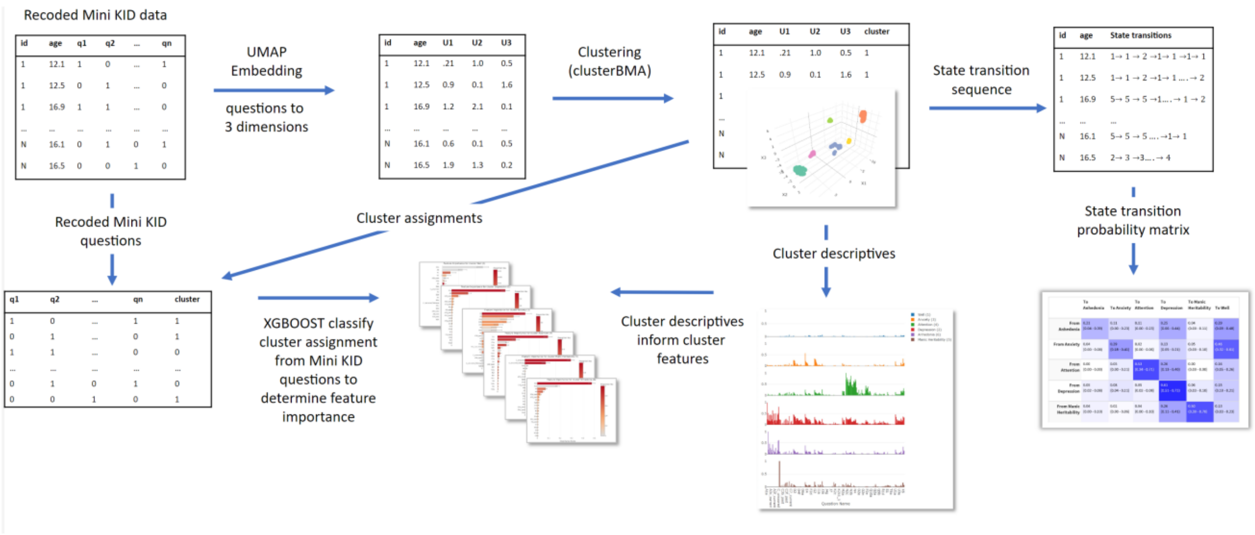


**Figure S2: Flowchart of the dimensionality reduction and cluster analyses.**

### Pluripotential Indexes (algorithm)

States were defined globally, and a state transition probability matrix was calculated using Markov chain modelling. We introduce a pluripotential index from the Shannon entropy of each row of the transition probability matrix. The maximum pluripotential occurs when there is equal probability of transitioning to any state in the matrix. The pluripotential index ($P_i$) for state $i$ is defined as:

$$P_i = -\frac{\sum_{j=1}^{k} p_{ij} \log(p_{ij})}{\log(k)} \quad \text{(Equation S1)}$$

where $p_{ij}$ is the probability of transitioning from state $i \rightarrow j$, and $k$ is the number of states. A value of zero indicates no transitions, while a value of one means equal probability of transitioning to any

### 2. Supplementary Results

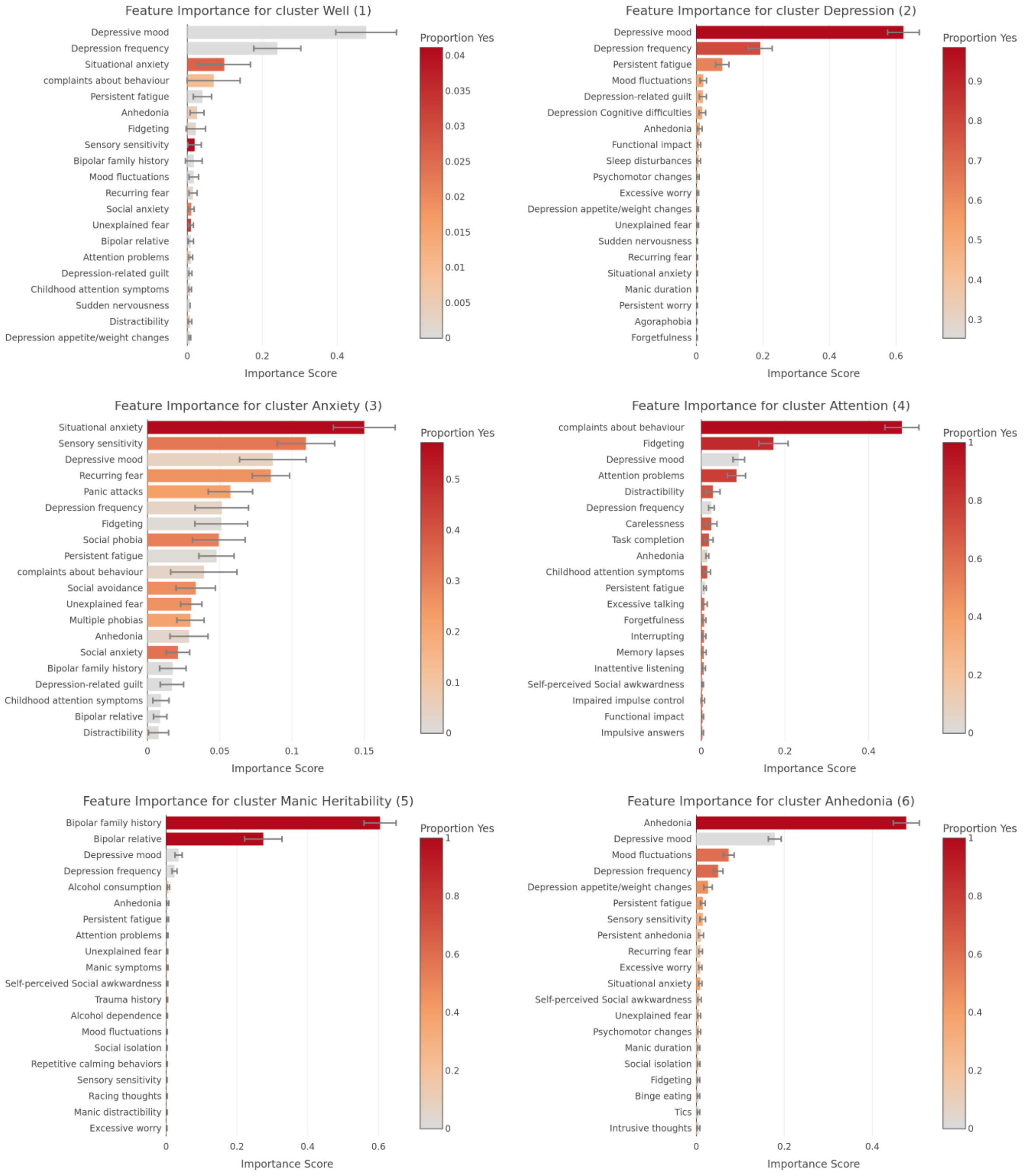


**Figure S3: Feature Importance for each state**

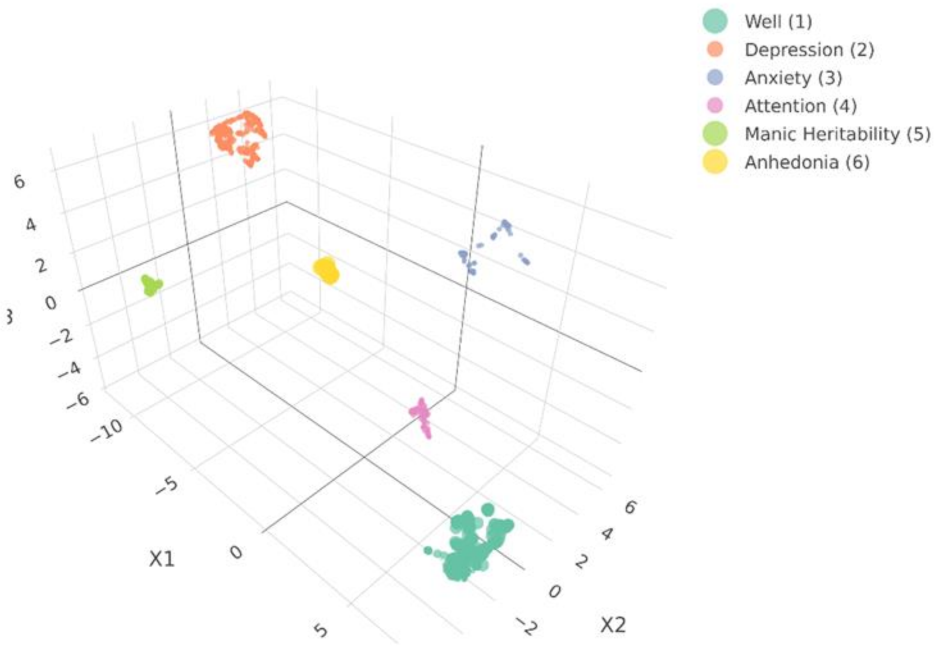


**Figure S4: UMAP embeddings labelled by state. Points represent observations, larger size points represent greater symptom allocation uncertainty.**

**Mental Health Seeking tables**

**Table S2: Mental health professional help seeking by age-group.**

| Age group | N | N help seeking | % help seeking |
|---|---|---|---|
| 12 | 85 | 41 | 48.2 |
| 13 | 113 | 66 | 58.4 |
| 14 | 104 | 60 | 57.7 |
| 15 | 80 | 51 | 63.8 |
| 16 | 52 | 28 | 53.8 |
| 17 | 38 | 18 | 47.4 |

**Table S3: Mental health professional help seeking at any time.**

| Help Seeking | N | % |
|---|---|---|
| FALSE | 70 | 42.2 |
| TRUE | 96 | 57.8 |

**Table S4: Fraction help seeking by age group and sex.**

| age group | sex | N | N help seeking | % help seeking |
|---|---|---|---|---|
| 12 | Female | 50 | 26 | 52.0 |
| 12 | Male | 35 | 15 | 42.9 |
| 13 | Female | 61 | 41 | 67.2 |
| 13 | Male | 52 | 25 | 48.1 |
| 14 | Female | 56 | 36 | 64.3 |
| 14 | Male | 48 | 24 | 50.0 |
| 15 | Female | 44 | 32 | 72.7 |
| 15 | Male | 36 | 19 | 52.8 |
| 16 | Female | 26 | 19 | 73.1 |
| 16 | Male | 26 | 9 | 34.6 |
| 17 | Female | 20 | 8 | 40.0 |
| 17 | Male | 18 | 10 | 55.6 |

**Table S5: Mental health professional help seeking at any time by sex.**

| Any help seeking | sex | N | % |
|---|---|---|---|
| FALSE | Female | 35 | 21.1 |
| | Male | 35 | 21.1 |
| TRUE | Female | 58 | 34.9 |

**Table S6: Mean psychological distress (K10 score) and Wellbeing (COMPAS-W Wellbeing score) by age group and state (95% CIs in brackets).**

| Age Group | Well (1) | Depression (2) | Anxiety (3) | Attention (4) | Manic Heritability (5) | Anhedonia (6) |
|---|---|---|---|---|---|---|
| **K10 score** | | | | | | |
| 12 | 12.8(12.1-13.4) | 20(16.9-23) | 17.8(15.1-20.4) | 15(11.4-18.6) | 16(10.4-21.6) | 17.1(14.4-19.8) |
| 13 | 13(12.4-13.6) | 24.7(20.8-28.7) | 15.7(14.1-17.4) | 15.8(13.8-17.7) | 16.7(12.7-20.7) | 17.2(12.8-21.6) |
| 14 | 13.8(13-14.7) | 23(20.4-25.5) | 16.5(13.8-19.2) | 19.3(16.7-21.8) | 15(13.2-16.8) | 22(18.5-25.5) |
| 15 | 14.2(13-15.4) | 22.5(19.6-25.4) | 17.9(14.1-21.7) | 18.5(14.4-22.6) | 16.6(13.6-19.6) | 20.9(15.3-26.4) |
| 16 | 14.4(13.2-15.6) | 25(21.3-28.6) | 16.4(13.7-19.1) | 20.5(17-24) | 14.6(12.3-16.9) | 15.8(13.6-17.9) |
| 17 | 14.4(13-15.9) | 25.2(21.4-28.9) | 16.3(13.7-18.8) | 18(15.2-20.8) | 14.9(13.2-16.6) | - |
| **COMPAS-W Wellbeing** | | | | | | |
| 12 | 105.7(102.8-108.6) | 97.4(93.1-101.8) | 101.1(96-106.1) | 101.7(95.1-108.2) | 105(99.4-110.6) | 93.4(85.7-101) |
| 13 | 105.1(103.1-107.2) | 91.5(85.8-97.1) | 98.5(94.7-102.3) | 96.6(92.4-100.7) | 101.8(95.1-108.5) | 100.8(95.6-106) |
| 14 | 104.2(101.6-106.8) | 91.1(86.4-95.7) | 100.3(95.7-105) | 88.3(83.2-93.4) | 101.3(96-106.6) | 91.8(84.5-99.1) |
| 15 | 103.9(100.8-107) | 90.5(86.4-94.7) | 96(87.6-104.4) | 92.4(84.4-100.5) | 102.2(97.9-106.5) | 97.4(90.3-104.6) |
| 16 | 105.8(102.6-109.1) | 90.6(86.5-94.8) | 100.2(96.9-103.5) | 86.5(67.7-105.3) | 106.1(100.2-112) | 107(103.1-110.9) |
| 17 | 104.4(100.1-108.6) | 90.7(84.4-96.9) | 98.3(90.2-106.3) | 79.5(73.2-85.8) | 108.3(105.8-110.8) | - |
